# Digital Nanofluidic Chip for Simple and Highly Quantitative Detection of HPV Target

**DOI:** 10.1101/2024.03.01.24303620

**Authors:** Li Liu, Stephen J. Dollery, Gregory J. Tobin, Guoyu Lu, Ke Du

## Abstract

Quantitative analysis of human papillomavirus (HPV)-infected cervical cancer is essential for early diagnosis and timely treatment of cervical cancer. Here, we introduce a novel energy transfer-labeled oligonucleotide probe to enhance the loop-mediated isothermal amplification (LAMP) assay for highly sensitive and specific detection of HPV 16. Conducted as a single-step assay within a digital nanofluidic chip featuring numerous reaction reservoirs, our method facilitates target amplification under isothermal conditions. Targeting an HPV 16 gene, our chip demonstrates the capability to detect HPV DNA at concentrations as low as 1 fM, spanning a dynamic range of five orders of magnitude. Importantly, our digital chip allows for highly quantitative detection of target genes at low concentrations, with the correlation between target concentration and the number of microwells exhibiting fluorescence signals. Furthermore, we have developed a computer vision method for automated and 100% accurate quantification of target concentrations. This research holds promising applications in clinical diagnosis and is poised for seamless integration into both hospital and point-of-care settings.

## Introduction

Cervical cancer is the fourth most common cancer in women globally, and in almost all instances this begins with Human papillomavirus (HPV) infection.^1^ The two most common high-risk genotypes HPV16 and 18 are responsible for over 70% of cervical cancer cases.^2^ Due to their elevated sensitivity and specificity, HPV DNA tests have become the gold standard for cervical cancer diagnosis.^3^ However, these PCR based DNA tests rely on bulky and expensive instruments such as thermal cyclers, making them impractical for point-of-care (POC) settings.^4^ In recent years, nucleic acid isothermal amplification detection has emerged as a promising alternative to traditional PCR due to its simplicity, speed, cost-effectiveness, and high sensitivity. Various nucleic acid isothermal amplification methods have been developed, including loop-mediated isothermal amplification (LAMP),^5^ recombinase polymerase amplification (RPA),^6^ rolling circle amplification (RCA),^7^ and nucleic acid sequence-based amplification (NASBA).^8^ Even though highly promising, most isothermal amplification detection methods have limitations such as a lack of quantitative detection capabilities, occasional unexpected non-specific amplification signals, and relatively high background signals.^9^

LAMP is one of the most widely applied isothermal amplification technologies in pathogen diagnostics.^10^ The advantages of the LAMP reaction include high sensitivity and the use of multiple primers, which have exceptionally high specificity because a set of four primers with six binding sites must hybridize correctly to their target sequence before DNA biosynthesis occurs.^11^ The reaction temperature is around 63°C, which avoids undesirable pre-amplification at room temperature.^12^ In addition, LAMP exhibits strong tolerance to inhibitors. Traditional detection methods relying on intercalating dyes like EvaGreen or SYBR Green can directly detect amplified targets but can have drawbacks such as low selectivity and the potential inhibition of the amplification reaction,^13^ thus requiring post-reaction analysis and causing false positives.^14^ To address these issues, energy transfer-labeled oligonucleotide probes for sequence-specific fluorescence detection have been developed to improve the amplification process. Here, we introduce an innovative ribonuclease-dependent cleavable FQ LB (fluorophore−quencher Loop B) primer designed to augment the sensitivity and specificity of LAMP detection. This primer undergoes cleavage exclusively upon recognizing a specific nucleic acid sequence, thereby markedly enhancing detection specificity and minimizing background signals.

Digital micro-and nanofluidic detection chips have been developed in recent years with advantages over traditional methods such as greater tolerance to inhibitory substances, higher sensitivity, and more accurate detection.^15^ The combination of digital chips and nucleic acid amplification methods enables absolute quantification analysis of nucleic acids targets by distributing target molecules into small wells or droplets. When performing limited dilution into aliquots, those aliquots contain no target molecule or only one molecule. The target molecule concentration can then be derived from counting the number of positive aliquots. The isolation of aliquots eliminates the competition of primers and probes, which is especially important for detecting minute DNA targets.^16^ Currently, there are mainly two strategies for generating isothermal reaction units, one of which involves using chamber microfluidic chips to generate reaction units. However, due to manufacturing process limitations, the number of chambers in microfluidic chips is limited, or the chamber volume is relatively large, leading to restricted dynamic range.^17^ The other strategy involves using droplet microfluidic chips to generate monodisperse droplets. However, the process of droplet formation is relatively complex, and the stability of the droplets poses issues, making real-time reaction monitoring a significant challenge.^18^

Here, we show a novel energy transfer-labeled oligonucleotide probe with improved fluorescence properties to create a highly sensitive and specific isothermal amplification of nucleic acids for sensitive and quantitative detection of HPV 16 sequences in plasmids which mimic dsDNA HPV genomes. The digital warm start assay is established through LAMP-based reaction in sub-microliter aliquots within a digital nanofluidic chip. This reaction is a one-pot format FQ labeled primer isothermal amplification-based detection, preventing premature target amplifications at room temperature and enabling accurate digital quantification of nucleic acids. Separation of aliquots eliminates competition between primers and probes and largely reduces false positive signals. Machine learning facilitates the straightforward derivation of quantitative relationships for target concentrations from the analysis. Our developed combination of the assay and nanofluidic chip allows for highly sensitive and uncomplicated detection of HPV 16. This system is poised for adaptation in hospitals or POC settings for the quantification of sexually transmitted infections.

## Experiments

### Materials and reagents

Deoxynucleotide (dNTP) solution mix (10 mM of each), Bst 2.0 WarmStart DNA polymerase (8 U/μL), MgSO_4_ (100 mM), 10 × Isothermal Amplification Buffer (200 mM Tris–HCl, 500 mM KCl, 100 mM (NH4)_2_SO_4_, and 20 mM MgSO_4_, 1.0% Tween 20 and pH 8.8 at 25 °C) were purchased from New England BioLabs (Ipswich, MA). Primers, FQ probe and the plasmid pCDNA3.1(+) containing 450-bp HPV 16 and HPV 18 gene sequences were purchased from either Integrated DNA Technologies (Coralville, IA) or synthesized by Genscript. All the sequence information of the primers and target sequences is listed in **Table S1**. The primer sequences were designed according to the LAMP Primer Design Tool from the New England BioLabs website. QuantStudio 3D digital 20K chip kit (Version 2) was purchased from Thermo Fisher Scientific (Waltham, MA).

### Preparation of the target plasmids

The 450-base pair (bp) target sequences of HPV-L1 gene from HPV16 and HPV18 were synthesized by GenScript (GenScript USA, Inc, Piscataway, NJ, USA) with HindIII and NotI restriction enzyme sites flanking the sequence. These restriction sites were incorporated to facilitate subsequent cloning steps. The synthesized gene was designed to yield a 463-bp product that was then inserted into the cDNA3.1(+) expression vector to yield a 5,820 bp plasmid. The insert sequence was verified post-synthesis through sequencing analysis (QuintaraBio, Frederick, MD, USA).

### Off-chip isothermal amplification assay

We performed all the HPV 16 target detection with FQ LB (fluorescence quenched) probe-based LAMP assay. Briefly, 25 μl of the reaction for each experiment contained 2.5 μl of 10 × Isothermal Amplification Buffer, 1.5 μl of MgSO_4_ (100 mM), 3.5 μl dNTP mix (10 mM), 2.5 μl of 10 × target-specific primer mix (FIP/BIP 16 μM, F3/B3 2 μM, LoopB 4 μM, FQ LB probe 4 μM), 1 μl of Bst 2.0 WarmStart DNA Polymerase (8 U/μl), and 2 μl of HPV target with different concentrations. The rest of the reaction contained 12 μl of nuclease-free water. The LAMP conditions were optimized for the reaction with a mixture of LB/FQ LB primers at concentrations of 0.2, 0.4, and 0.6 μM, respectively. We also optimized the ratio of LB/FQ LB primers with 2:1, 2:2, and 2:4, respectively. The reaction was incubated at 63°C for 45 min. After incubation, the tubes were placed under a Maestrogen UltraSlim LED blue light illuminator (Pittsburgh, PA) to take images (Canon EOS 6D, Canon, Japan). The endpoint fluorescence was measured by a plate reader (BioTek Cytation 5, Agilent, CA, USA).

### On-chip isothermal digital amplification assay

The digital nanofluidic chip is a 10 mm^2^ high-density reaction plate that has a single array of 20,000 reaction microwells with 15 μl volume for each well. The 15 μl reaction mixture contained 1.5 μl of 10 × Isothermal Amplification Buffer, 0.9 μl of MgSO_4_ (100 mM), 2.1 μl dNTP mix (10 mM), 1.5 μl of 10× target-specific primer mix (FIP/BIP 16 μM, F3/B3 2 μM, LoopB 4 μM, FQ LB probe 4 μM), 0.6 μl of Bst 2.0 WarmStart DNA Polymerase (8000 U/ml), 1.2 μl of target DNA, and 7.2 μl of nuclease-free water. After assay mixing, the 15 μl reaction mixture was loaded into the chip and mineral oil was used to seal the loading port. The device was placed on a heat block and incubate at 63°C for 45 min.

### Data acquisition and analysis

After incubation, the chip was quantified using a BioTek Cytation 5 Cell Imaging Multimode Reader (Agilent, CA, USA) with a 4× magnification objective. Every image was captured in ∼30 s of laser irradiation, and the irradiation was turned off until the next imaging. Five distinct regions (3.5 × 3.5 mm) without overlapping areas were randomly captured by the microscopy to cover about 2,750 microwells. The number of the positive spots and corresponding fluorescence intensity were measured by using the ImageJ software (bit depth: 8 bits; 1,992 × 1,992 pixels). GraphPad Software Prism 9.5.1 was used to plot real-time fluorescence curves, analyze linear regression, and verify statistical significance between different assay groups.

## Results

The schematic of the digital nanofluidic chip based on the one-step LAMP assay is shown in **Figure 1a**. The primers were designed to amplify an HPV 16 L1 fragment gene sequence (GenBank accession MT316211.1). The FQ-LAMP reaction mixture was first prepared in an Eppendorf tube. The prepared reaction mixture was then distributed into a QuantStudio 3D digital chip. The 3D digital chip is a 10 mm^2^ high-density reaction plate that has a single array of 20,000 reaction microwells. Each microwell has a diameter of 60 μm and a depth of 500 μm (**Figure 1b**). The chip was pretreated with hydrophobic coating on the chip surface to enable the loading and isolation of the LAMP reactions within the microwells. After the addition of the reaction reagent, an oil layer was applied to cover the chip. This step not only facilitates the straightforward isolation of each reaction microwell but also serves as a preventive measure against contamination and reagent evaporation. Following a 45-minute incubation at 63°C, microwells containing the target DNA exhibit a green fluorescence signal attributed to successful target amplification, while microwells lacking the target remain devoid of such fluorescence.

**Figure 1.**
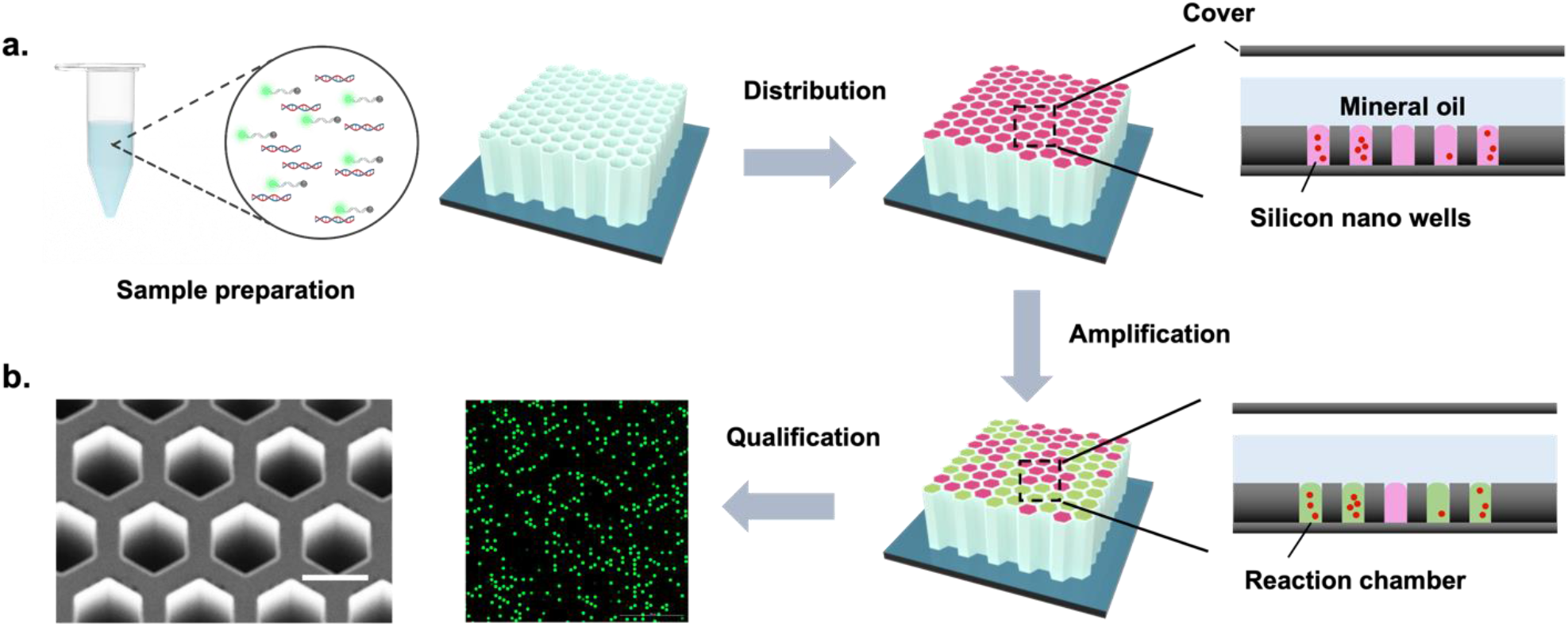
(a) Schematic illustration of FQ LAMP assay. One-pot reaction mixture is first prepared in one tube. The sample mixture then distributed randomly into over ten thousand of microwells. When incubated at 63 °C, each micro reaction with plasmid HPV 16 DNA target undertakes reaction and is amplified, generating strong green fluorescence (positive spots), whereas not in those without target (negative spots). Through detecting and counting the positive micro reactions (or spots), plasmid HPV 16 DNA can be quantified based on the proportion of positive spots. (b) SEM image of micro digital chip, Scale bar is 60 μm.

**Figure 2a** shows the schematic of the FQ probe-based LAMP reaction. This assay was developed with the designation of two sets of primers to specifically detect the HPV 16 DNA target. Five LAMP primers (FIP, BIP, F3, B3, and LB) were designed according to the distinct regions of the HPV 16. In the present study, two different types of LBs were constructed: the LB probe is the traditional probe with unlabeled ends. The other probe, named the FQ LB probe, has the same sequence as the LB probe but was tagged with FAM fluorophore at the 5′-end and Iowa Black® RQ quencher at the 3′-end. This probe is quenched at the unbound state and fluoresces only when annealed to the specific complementary regions during the amplification process. The quencher functions to inhibit the fluorophore from emitting signals when they are close to each other. The fluorophore and quencher were placed further from each other to allow it specifically annealing the stem loop region of the dumbbell like LAMP amplicons. As the probe is longer than 20 base pairs, it was designed with an additional internal quencher, that is, an internal quencher /ZEN/ positioned in the middle of the strand. This design was intended to reduce the assay’s crosstalk signals, increase the amplification signal, and produce a lower background noise. As shown in **Figure 2b** and **Figure 2c**, the optimal concentration of the LB and FQ LB primers and the optimal ratio of LB primer to FQ LB primer are 0.4 μM and 1:1, respectively. The fluorescence intensity was measured for the target DNA concentration ranging from 10 aM to 1 pM (**Figure 2d**), and the positive reaction was clearly observed in the 1 fM sample with both naked eye and UV illumination. **Figure 2e** shows the comparison of the fluorescence intensity for positive and negative targets (1 pM), and a clear difference between positive (HPV16) and negative groups (HPV 18 and NTC) is observed.

**Figure 2.**
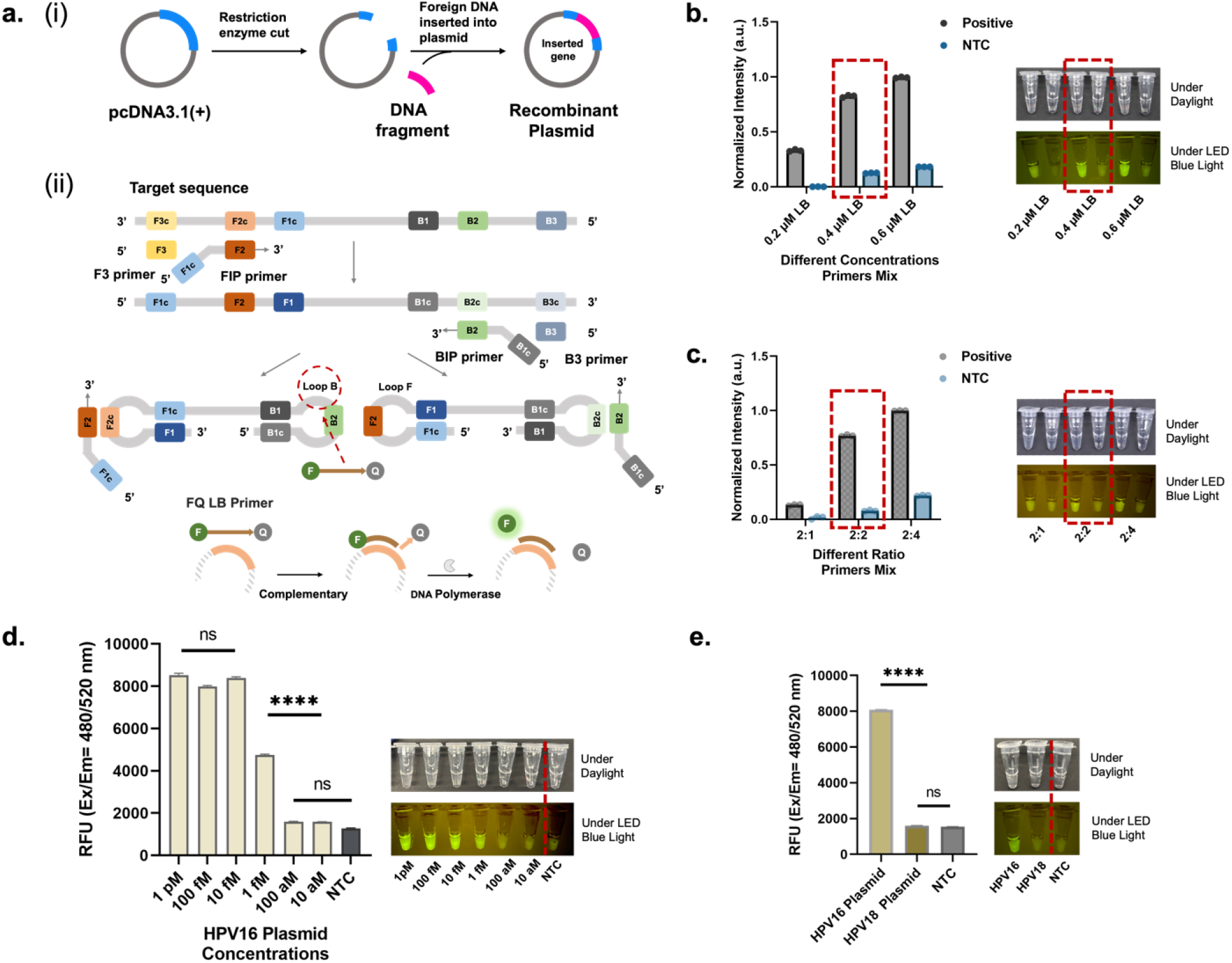
(a-i) Target sequences of HPV-L1 gene from HPV16 insert into plasmid cDNA3.1(+) vector (450 base pairs). (a-ii) Principle of the FQ-LAMP assay. In the assay, LAMP’s loop backward (LB) primer was used to design the FQ LAMP probe. (b) The LAMP conditions were optimized for the reaction with a mixture of LB/FQ LB primers at concentrations of 0.2, 0.4, and 0.6 μM, respectively. (c) The LAMP conditions were optimized for the reaction with a mixture ratio of LB/FQ LB primers were 2:1, 2:2, and 2:4, respectively. (d) Endpoint fluorescence detection of the FQ LAMP after 45-min amplification of various concentrations of plasmid HPV 16 DNA targets, from 10 aM to 1 pM. (e) Endpoint fluorescence detection of the Plasmid HPV 16 DNA target (1 pM) compared with plasmid HPV 18 DNA target (1 pM) and no template control. For each concentration’s testing, error bars denote the standard deviation (n = 3). The asterisks represent statistical significance according to a t-test. *P ≤ 0.05, **P ≤ 0.01, ***P ≤ 0.001, ****P ≤ 0.0001.

After optimizing the reaction conditions off-chip, the analytical performance of the microwell chip was investigated. **Figure 3a** illustrates the typical workflow of the FQ-LAMP assay, encompassing the synthesis of DNA Plasmid which mimics the HPV circular dsDNA genome, preparation of a one-pot LAMP reaction mixture, distribution of the reaction mixture into the chip, and on-chip incubation at 63°C. **Figure 3b** shows the images of on chip reactions under a microscope with various incubation times (e.g., 0, 15, 30, 45, 60 and 90 min) using 1 pM HPV plasmid as a target. As shown in **Figure 3c**, a 45-min incubation is enough for the FQ probe LAMP assay to reach the maximum percentage of the positive spots. The uniformity of the microwell volume was also characterized by ImageJ. As shown in **Figure 3d**, the average well area was 790.7 ± 9.633 Pixel^2^, which conforms to a Gaussian distribution (R^2^=0.9805).

**Figure 3.**
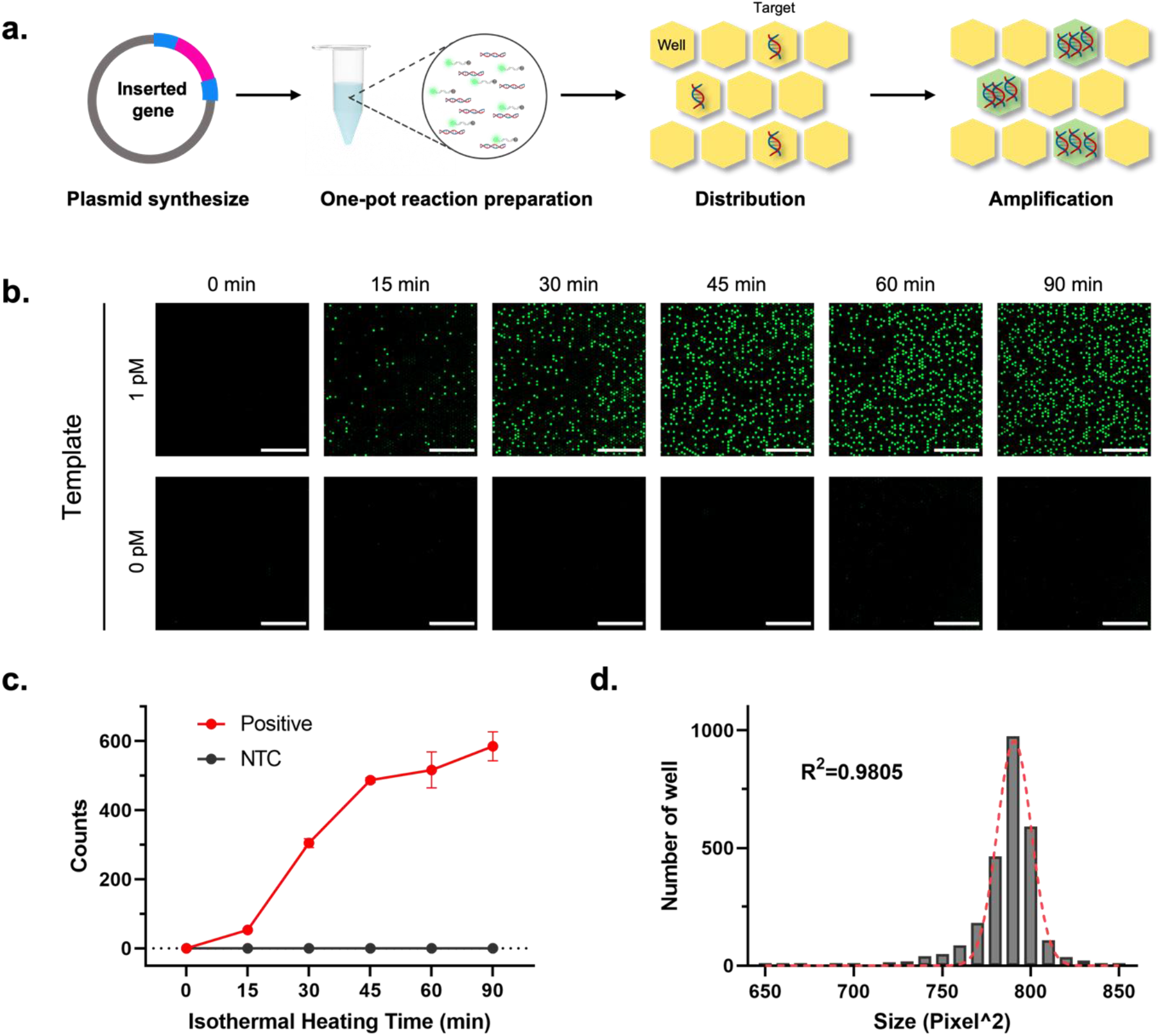
(a) The schematic of digital amplification in microwells. (b) Endpoint fluorescence micrographs of the digital chip for the HPV 16 detection (1 pM) with various incubation time (0, 15, 30, 45, 60 and 90 min) at 63 °C. (c) The average positive spots increase to a detectable level within 15 min, and when the time was greater than 45 min, there was no significant change in the positive spots. Error bars denote the standard deviation (n = 5). (d) Histogram of well area conforming to Gaussian distribution (R^2^=0.9805), proofing the high uniformity of the reagents in the microwells.

By testing various concentrations of HPV 16 DNA plasmid target, the on-chip detection sensitivity was also investigated. As shown in **Figure 4a**, the digital FQ LAMP assay was performed with a target concentration ranging from 100 aM to 100 pM, and signals can directly be observed by counting the positive spots shown under the microscope. We are able to detect the HPV 16 target with a concentration as low as 1 fM, and the signal is saturated with a concentration greater than 10 pM. The specificity of the digital LAMP assay was also carried out by using similar strains. As shown in **Figure 4c**, positive spots are observed in the chip loaded with the HPV 16 positive control, whereas not for those negative control samples, such as the HPV 18 control and the no template control, which is consistent with our off-chip results. In addition, the digital nanofluidic chip shows highly quantitative readings with a target concentration range from 1 fM to 10 pM (**Figure 4b**), demonstrating a dynamic range of 5 orders of magnitude. Moreover, notable distinctions exist in the specificity of HPV target detection when compared to other strains (**Figure 4d**).

**Figure 4.**
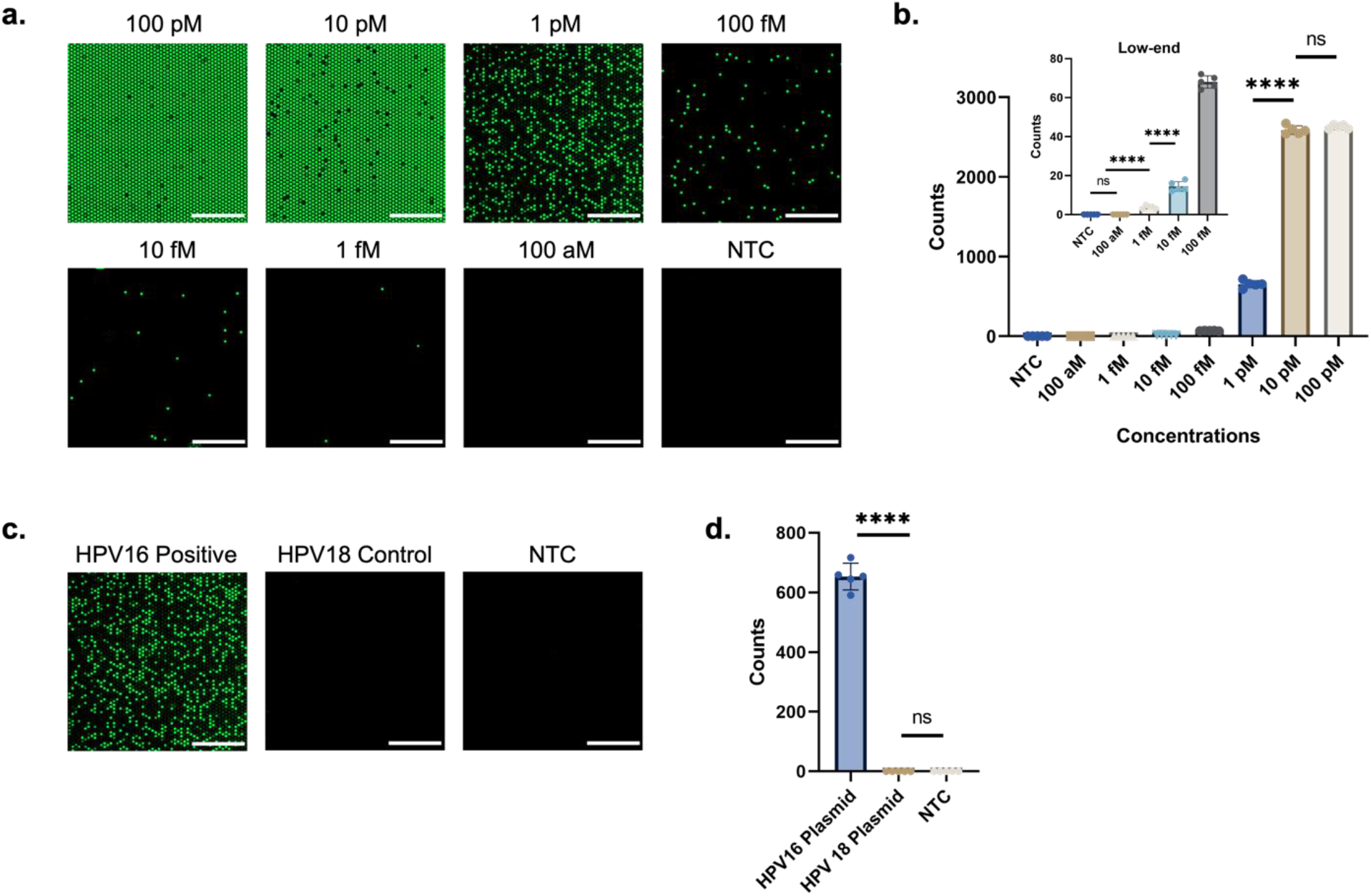
(a) End-point fluorescence images of reactions with different starting concentrations of plasmid HPV 16 DNA target within 45 min incubation at 63 °C. Scale bar, 1 mm. (b) Quantification range of the digital chip. The relationship between positive spots (Y) and concentration of targets (X), from 100 aM to 100 pM. The low-end shows the enlarged view of low concentration range from 100 aM to 100 fM. (c) End-point fluorescence images for the specificity of Plasmid HPV 16 PC, Plasmid HPV 18 control, and no template control. Scale bar: 1 mm. (d) Detection specificity of the Plasmid HPV 16 DNA target (1 pM) compared with plasmid HPV 18 DNA target (1 pM) and no template control. For each concentration’s testing, error bars denote the standard deviation (n = 5). The asterisks represent statistical significance according to a t-test. *P ≤ 0.05, **P ≤ 0.01, ***P ≤ 0.001, ****P ≤ 0.0001.

The entire pipeline for processing images and classifying the testing samples is shown in **Figure 5**. To estimate target concentrations from sample images, we first inputted a dataset of training images showcasing results. These images are crucial for creating a baseline for the system’s understanding of target appearances at various concentrations. Next, template matching algorithms were applied to detect samples within these images due to the spatial hexagon shape property of each individual sample. We slide this hexagon template across the entire sample image to detect individual samples. This precise detection is critical for accurate analysis. The detected samples were sorted based on image intensity levels, arranging them from high intensity to low intensity.

**Figure 5.**
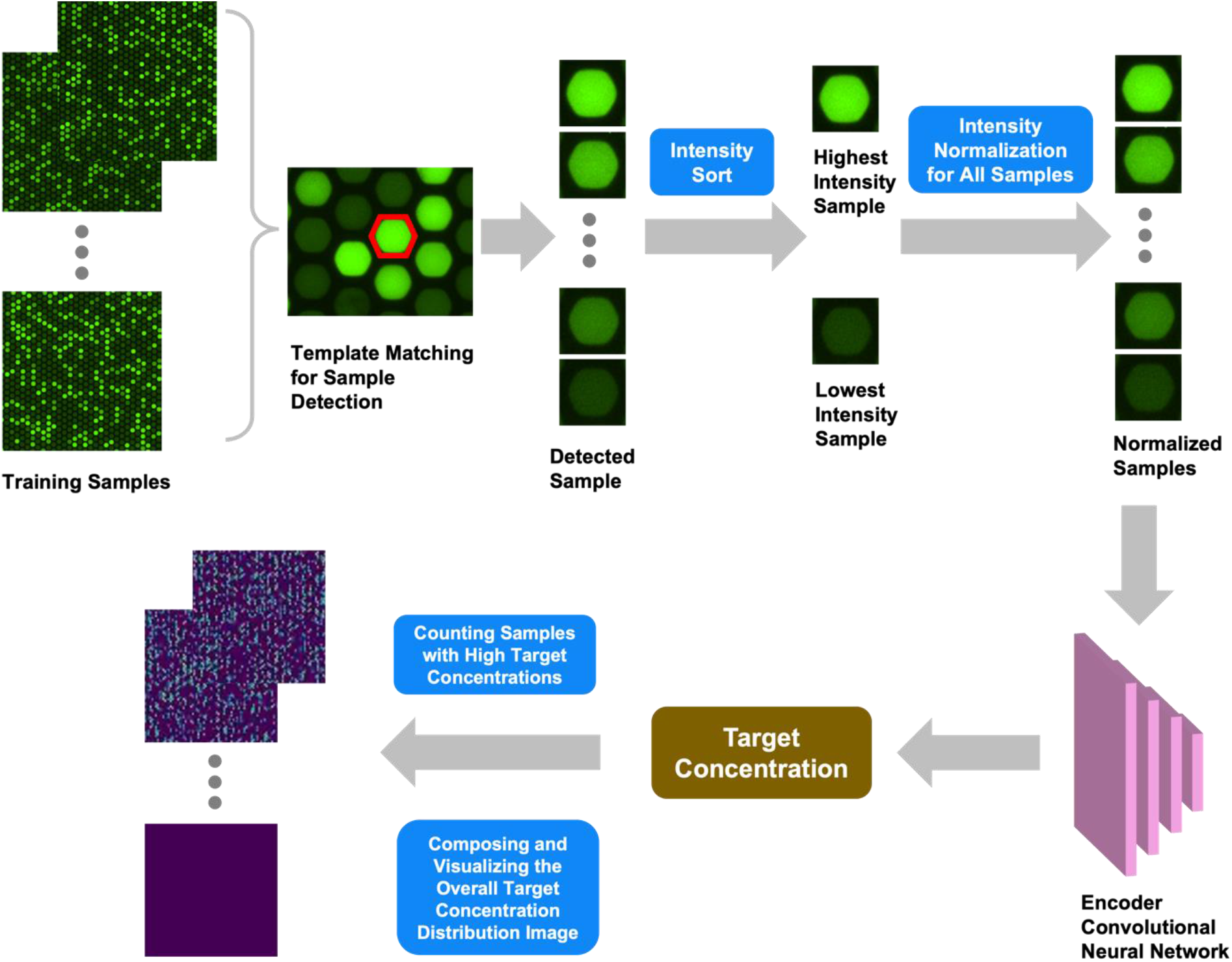
Image processing and testing result classification pipeline of computer vision enabled HPV target sensing on Digital Nanofluidic Chip, which involves feature generation, intensity range selection, and feature classification.

This sorting is essential to standardizing the analysis across different samples, as the sample image intensity is highly correlated to the target concentration. Subsequently, the intensity values of all samples were normalized to account for variations in target concentration. This normalization ensures consistent and reliable data fed into the neural network. Finally, a Convolutional Neural Network (CNN) was trained using these normalized images. The CNN learns the patterns and correlations between the visual characteristics of the target samples and their known concentrations. This learning enables the network to accurately estimate target concentrations in new, unseen images. Eventually, we composed the target concentration result for each individual sample to entire training sample image with thousands of individual samples and visualize the target concentration distribution. With this learning algorithm, our system can correctly identify all the positive and negative samples, and the network can accurately estimate the target concentration in unseen images.

## Discussion

In this study, we developed a novel energy transfer-labeled oligonucleotide probe to create a highly sensitive and specific isothermal amplification assay for nucleic acids detection. We applied it in a digital nanofluidic chip to enable rapid quantitative detection of HPV targets. FQ LB is a single fluorophore and quencher-labeled oligonucleotide probe that can be cleaved by ribonuclease, simultaneously initiating rapid nucleic acid amplification and generating sequence-specific fluorescence. The devised assay is conducted as a single-step within a closed one-pot procedure, mitigating the risk of carryover or cross-contamination arising from post-amplification procedures. The quencher restricts the signals by the fluorophore when they are in close proximity. The probes employed in our approach are quenched in the unbound state and emit fluorescence exclusively upon annealing to a specific complementary region during the amplification process, ensuring high sequence specificity. For LB probes shorter than 20 nucleotides, labeling is limited to the ends, while those exceeding 20 nucleotides incorporate an internal quencher at the midpoint, which would otherwise not be quenched due to the length of the DNA strand. This design reduces crosstalk, enhances amplification, and diminishes background noise.^19^ In addition, the use of FQ probes does not require a complex pre-processing procedure. Due to the complexity of distinguishing non-specific LAMP amplicons from target amplicons, FQ probes are more specific than conventional LAMP assays using DNA intercalating dyes and can significantly reduce background signals. Therefore, this study reveals the benefits of using FQ LB probes to improve amplicon detection in LAMP assays.

We developed a robust digital amplification system for easy identification of detection targets. This method is a simple patterning method that is unique in its ability to concentrate reagents in a small size. Because the target molecules are assigned to numerous small, separated reaction reservoirs, the positive and negative reservoirs can easily be distinguished from each other without interference. The system is more tolerant to reaction inhibitors, as the potential reaction inhibitors are separated from the reaction mixture, reducing the amplification reaction inhibition in the digital assay and making it well suited for the detection of low-level targets. We found that due to the multiprobing of the conventional LAMP assays, reactions in Eppendorf tubes are more susceptible to non-specific amplifications due to the cross-linking of amplicons, and aerosols generated during the reaction may cause contamination leading to false positives.^20^ In contrast, no false-positive signals were found in the reaction system of our digital nanofluidic chip. Therefore, digital detection can directly detect nucleic acid in many samples without complicated sample pre-treatment and nucleic acid purification processes.

Our microliter digital microwells enable the quantitative detection of nucleic acids without the need of calibration curves, thereby improving the accuracy of detecting low-copy nucleic acid template. We found that conventional reactions in Eppendorf tubes are difficult to achieve accurate quantitative detection and are more suitable for qualitative detection. In contrast, the microliter digital microarray reaction allows for easy quantitative distinguish across five orders of magnitude, ranging from 1 fM to 10 pM. The ability to quantitatively detect nucleic acids without relying on calibration curves is a key feature that enhances accuracy, particularly in low-copy nucleic acid template detection. In addition, compared to droplet microfluidic chips that produce monodisperse droplets, fixed-structure chambers are more stable and suitable for real-time monitoring. The compartmentalization of nucleic acid in digital detection enables individual amplification and detection, enhancing sensitivity to the single-molecule level. This surpasses the sensitivity achievable with traditional isothermal amplification reactions, which often require intricate primer screening and probe optimization. Currently, we have labeled different targets with different fluorophores and quenchers, allowing them to be detected simultaneously on different fluorescent channels of a real-time monitoring system.^21^

We have developed a computer vision-based data analysis system. Through this learning process, our system correctly identifies both positive and negative samples, and the network can also accurately estimate the concentration of targets in unseen images. In the end, the classification accuracy of separating detected positive samples from negative samples is 100%. Our method integrates image processing and deep learning techniques, making it a robust and powerful tool for analyzing target concentrations, which has important applications in areas such as molecular detection and clinical diagnosis. The application of computer vision in biosensors has great prospects, as it enables large-scale, automated, high-throughput, and multi-target detection compared with the traditional analysis methods. The prospect of enhancing sensitivity, specificity, and efficiency in biosensing through computer vision signifies a significant leap forward in the realm of diagnostic and analytical methodologies. It stands as a vital tool for the future development of traditional biosensors toward intelligent biosensors.^22^

## Data Availability

All data produced in the present study are available upon reasonable request to the authors

## Acknowledgments

This work was supported by NIH R35GM 142763, NIH 75N93023C00056-0-9999-1, and USDA USDA NIFA 2022-67021-41478.

## Conflict of interests

The authors declare that they have no conflict of interest.

## Notes

### Competing Interest Statement

The authors have declared no competing interest.

### Funding Statement

NIH R35GM 142763
NIH 75N93023C00056-0-9999-1
USDA USDA NIFA 2022-67021-41478

